# Predicting chemoresponsiveness in epithelial ovarian cancer patients using circulating small extracellular vesicle-derived plasma gelsolin

**DOI:** 10.1101/2022.10.13.22281057

**Authors:** Emma Gerber, Meshach Asare-Werehene, Arkadiy Reunov, Dylan Burger, Tien Le, Euridice Carmona, Anne-Marie Mes-Masson, Benjamin K. Tsang

## Abstract

**Background:** Resistance to chemotherapy continues to be a challenge when treating epithelial ovarian cancer (EOC), contributing to low patient survival rates. While CA125, the conventional EOC biomarker, has been useful in monitoring patients’ response to therapy, there are no biomarkers used to predict treatment response prior to chemotherapy. Previous work *in vitro* showed that plasma gelsolin (pGSN) is highly expressed in chemoresistant EOC cell lines, where it is secreted in small extracellular vesicles (sEVs). Whether sEVs from tumour cells are secreted into the circulation of EOC patients and could be used to predict patient chemoresponsiveness is yet to be determined. This study aims to determine if sEV-pGSN in the circulation could be a predictive biomarker for chemoresistance in EOC.

**Methods:** Sandwich ELISA was used to measure pGSN concentrations from plasma samples of 96 EOC patients (primarily high grade serous EOC). sEVs were isolated using ExoQuick ULTRA and characterized using western blot, nanoparticle tracking analysis, and electron microscopy after which pGSN was measured from the sEVs. Patients were stratified as platinum sensitive or resistant groups based on first progression free interval (PFI) of 6 or 12 months.

**Results:** Total circulating pGSN was significantly decreased and sEV-pGSN increased in patients with a PFI ≤ 12 months (chemoresistant) compared to those with a PFI > 12 months (chemosensitive). The ratio of total pGSN to sEV-pGSN further differentiated these groups and was a strong predictive marker for chemoresistance (sensitivity: 73.91%, specificity: 72.46%). Predetermined CA125 was not different between chemosensitive and chemoresistant groups and was not predictive of chemoresponsiveness prior to treatment. When CA125 was combined with the ratio of total pGSN/sEV-pGSN, it was a significant predictor of chemoresponsiveness, but the test performance was not as robust as the total pGSN/sEV-pGSN alone.

**Conclusions:** Total pGSN/sEV-pGSN was the best predictor of chemoresponsiveness prior to treatment, outperforming the individual biomarkers (CA125, total pGSN, and sEV-pGSN). This multianalyte predictor of chemoresponsiveness could help to inform physicians’ treatment and follow up plan at the time of EOC diagnosis, thus improving patients’ outcomes.

## Introduction

Despite epithelial ovarian cancer (EOC) being one of the most common gynecological cancers (1,2), late-stage presentation and chemoresistance present significant challenges to tumour control and oncologic outcomes. This ultimately results in an elevated case fatality rate among most EOC patients (1). Standard of care management of EOC includes a combination of aggressive surgical debulking and combination chemotherapy with a platinum drug and taxane derivatives either in a neoadjuvant and/or adjuvant clinical setting (3). Unfortunately, many patients will eventually recur due to development of chemoresistance (4). Chemoresistance is commonly defined in terms of progression free interval (PFI; time between completion of adjuvant chemotherapy and signs /symptoms of recurrent disease). Clinically, a PFI of 6 or 12 months is used as a cut-off to determine different degrees of platinum sensitivity (4). Presently, cancer antigen 125 (CA125) is the most commonly used biomarker in EOC to aid in the diagnosis, prognostication, and assessment of therapy effectiveness (5–7). While serum CA125 concentrations during and after chemotherapy are effective in monitoring disease response or progression, CA125 has not been shown to be effective in predicting response to chemotherapy prior to treatment (8). As such, there are no existing biomarkers that can be used to predict chemoresponsiveness prior to chemotherapy initiation.

Small extracellular vesicles (sEVs) are a subset of extracellular vesicles that range in size from ∼30 - 150nm (9). These small vesicles are released by all cell types and carry a molecular signature that reflects that of the originating cell (10), including nucleic acids, proteins, lipids, and metabolites (9). Because of their cargo and presence in circulation, sEVs are a promising source of minimally invasive biomarkers, as they can be retrieved from blood or urine samples.

In the context of ovarian cancer chemoresistance, many studies have identified cargo of extracellular vehicles (EVs) that play a role at the level of the tumour microenvironment to promote subsequent drug resistance (11–14). Studies are necessary to evaluate whether these EV cargo at the systemic circulatory level could be used to predict chemoresistance in EOC patients prior to the start of chemotherapy.

Gelsolin (GSN) is a calcium modulated actin-binding protein, playing an important role in cytoskeletal rearrangement, motility, and morphology (15). GSN has two well-studied isoforms; cytosolic GSN (cGSN) remains within the cell, while plasma GSN (pGSN) is the secreted isoform. These isoforms arise from different transcription start-sites and alternative splicing (15,16). pGSN plays an important role as an actin scavenger in the blood, preventing actin polymerization (17). Much work has been done to elucidate what role pGSN plays in resistance to chemotherapy in EOC. More specifically, pGSN within the tumour downregulates the anti-tumour functions of immune cells in the tumor microenvironment (CD8+ T cells, CD4+ T cells, dendritic cells and M1 macrophages) (18–20). pGSN is over-expressed in chemoresistant cells, transported via sEVs and confers resistance in otherwise chemosensitive cells (14). Although circulatory pGSN is indicative of early stage EOC and residual disease (21), its clinical utility in predicting chemoresistance is yet to be studied. Additionally, we have yet to examine the presence and clinical utility of sEV-derived pGSN (sEV-pGSN) in EOC.

In this study, we investigated whether sEV-pGSN presents as a better predictive biomarker of chemoresistance compared to total pGSN and CA125 prior to treatment. Identifying sEV-pGSN as an important predictor of chemoresistance would provide useful clinical information that could inform physicians’ treatment plan, follow up, and hopefully improve patient outcomes.

## Results

### Patient characteristics

This study used plasma samples from 96 EOC patients. Most of these patients had high-grade serous pathology (72%), while 26% of them have a non-serous subtype. One individual in this group had low-grade serous EOC. Further, approximately 70% of the samples were collected from individuals with FIGO stage III EOC. Few patients (9%) had recurrence within 6 months of treatment, while 24% had recurrence within 12 months. Further details of patient demographics are described in Table 1.

### Chemoresistant patients have elevated total pGSN but decreased sEV-pGSN at the time of diagnosis compared to chemosensitive patients

Although CA125 has been shown to be useful in monitoring EOC patients during and after chemotherapy (6,7), there is yet to be a validated biomarker that has a clinical application in predicting chemoresponsiveness *before* treatment initiation (22,23). In this cohort of patients, blood samples were collected prior to surgical debulking and chemotherapy. Predetermined CA125 was correlated with the patients’ response to chemotherapy treatment. Here, no difference in CA125 is observed between platinum resistant/partially resistant (PFI ≤ 6 or 12 months) and platinum sensitive/partially sensitive (PFI > 6 or 12 months) disease (Supplementary Figure 1A, Figure 1A).

**Figure 1.**
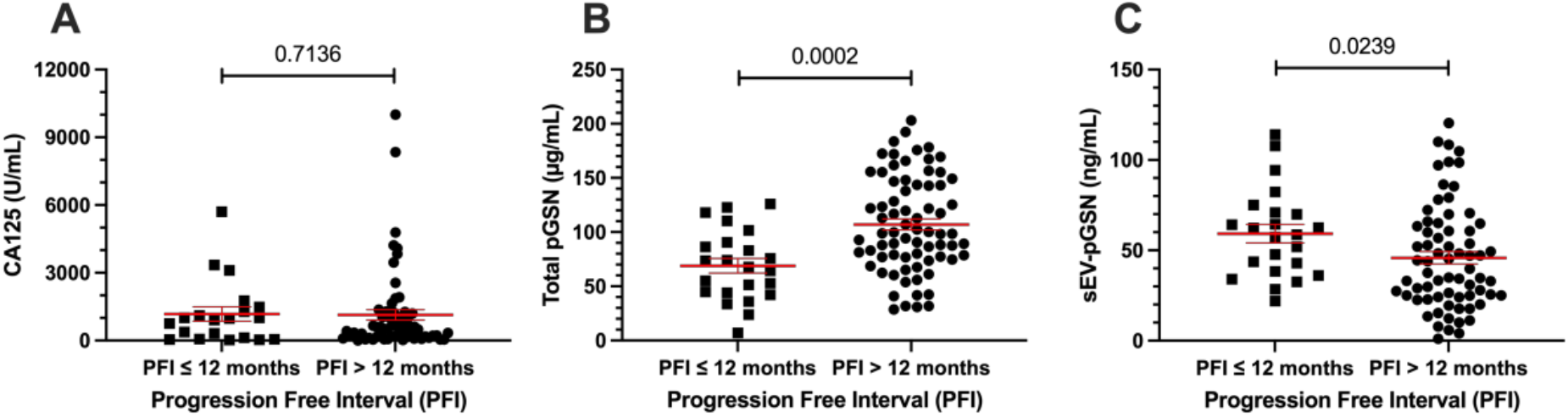
Total pGSN and sEV-pGSN are significantly associated with chemoresistance. Distribution of individual biomarkers between chemoresistant (PFI ≤ 12 months) and chemosensitive (PFI > 12 months) groups using dot plots. Points on dot plots represent individual patient biomarker concentrations. Line with error bars represent group mean and SEM. **(A)** CA125, Mann-Whitney U-test. **(B)** Total pGSN, Student t-test. **(C)** sEV-pGSN, Mann-Whitney U-test.

In the same group, total pGSN and sEV-pGSN were measured in the plasma samples. The mean level of pGSN was lower in patients with chemoresistance (PFI ≤ 12 months; 69 µg/mL ± 6.8) compared to chemosensitive patients (pGSN > 12 months; 107 µg/mL ± 5.2) (p = 0.0002, Figure 1B). Conversely, chemoresistant patients (PFI ≤ 12 months, 59 ng/mL ± 5.1) had elevated sEV-pGSN compared to chemosensitive patients (PFI > 12 months, 45.9 ng/mL ± 3.5) (p = 0.0239, Figure 1C). Although a similar trend was observed when patients were stratified by a PFI ≤ 6 months, the differences were not significant (Supplementary Figure 1B-C). These results suggest that total and sEV-pGSN have a potential clinical utility as biomarkers of chemoresistance (PFI ≤ 12 months) in EOC patients.

### The ratio of total pGSN/sEV-pGSN outperforms individual markers in predicting EOC chemoresistance

Previous studies in cancer and other diseases have demonstrated that multi-analyte panels of biomarkers outperform individual biomarkers in patient diagnosis and disease management (24–26). We investigated whether combining total pGSN with sEV-pGSN would enhance prediction of chemoresponsiveness in EOC patients compared to the individual markers. To do so, we calculated their ratio (total pGSN/sEV-pGSN) and compared the means (or mean ranks) between the groups. We found that regardless of PFI stratification chemoresistant patients had significantly lower total pGSN/sEV-pGSN (PFI, 6 months: p = 0.0264; PFI, 12 months: p < 0.0001) (Supplementary Figure 2A and Figure 2A). In comparison, neither individual marker showed a significant difference for PFI of 6 months (Supplementary Figure 1B-C). Taken together, these findings highlight the clinical importance of total pGSN/sEV-pGSN as a multi-analyte biomarker in differentiating chemoresistant EOC patients from chemosensitive patients, regardless of PFI stratification.

**Figure 2.**
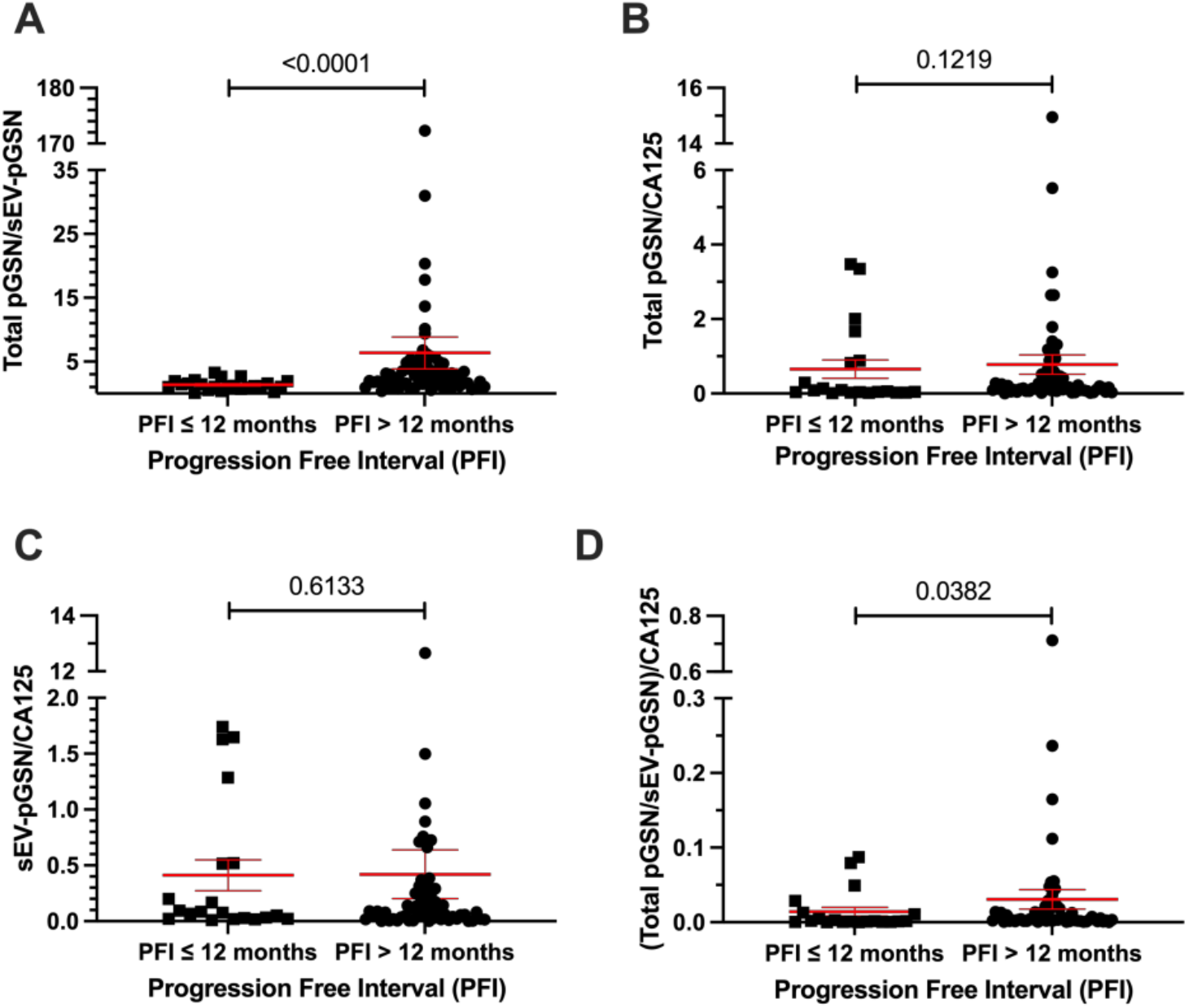
Total pGSN/sEV-pGSN shows the strongest association with chemoresistance. Distribution of multi-analyte biomarkers between chemoresistant (PFI ≤ 12 months) and chemosensitive (PFI > 12 months) groups using dot plots. Points on dot plots represent individual patient biomarker concentrations. Line with error bars represent group mean and SEM. **(A)** Total pGSN/sEV-pGSN. **(B)** Total pGSN/CA125. **(C)** sEV-pGSN/CA125. **(D)** (Total pGSN/sEV-pGSN)/CA125. Mann-Whitney U-test used for all four multi-analyte biomarkers.

### CA125 has no significant clinical utility in a multianalyte panel when differentiating chemoresistant from chemosensitive EOC patients

To investigate if using CA125 in a multi-analyte panel of biomarkers would further enhance the differentiation of chemosensitive from chemoresistant groups, we calculated two ratios using CA125 (total pGSN/CA125 and sEV-pGSN/CA125). Neither ratio had significant differences between chemosensitive and chemoresistant patients (PFI 6 months: Supplementary Figure 2B-C, PFI 12 months: Figure 2B-C). When the three biomarkers were combined by dividing total pGSN/sEV-pGSN by CA125, a significant difference was observed at both PFI 6months and 12 months (p = 0.0446 and 0.0382, respectively). Although a significant difference was observed with total pGSN/sEV-pGSN/CA125, this was not as strong as total pGSN/sEV-pGSN alone (without CA25), suggesting that CA125 adds no clinical value to total pGSN/sEV-pGSN in differentiating chemoresistant from chemosensitive EOC patients.

### Total pGSN/sEV-pGSN is the best biomarker combination to predict chemoresistance in EOC patients

We further examined the clinical test performance of the individual biomarkers and their combinations using receiver characteristic operating (ROC) curve analysis. In this cohort of patients, CA125 was unable to predict which individuals would have recurrence within 6 or 12 months of treatment (Supplemental Figure 3B and Figure 3B, respectively). Meanwhile, total pGSN predicted a PFI of 12 months with a sensitivity of 73.1% and specificity of 65.75% (cutoff = 86.37 µg/mL, AUC = 0.7451, p = 0.0004) and sEV-pGSN with a sensitivity of 60.87% and specificity of 62.32% (cutoff = 50.57 ng/mL, AUC = 0.6572, p = 0.0245) (Figure 3A). This was, however, not the case for PFI of 6 months (Supplemental Figure 3A). Most impressively, total pGSN/sEV-pGSN greatly improved the specificity of the test for predicting chemoresistance irrespective of the PFI stratification (Supplemental Figure 3A and Figure 3A). We found that this ratio could predict a PFI of ≤ 12 months with a sensitivity of 73.91% and a specificity of 72.46% (cutoff: 1.586, Figure 3A).

**Figure 3.**
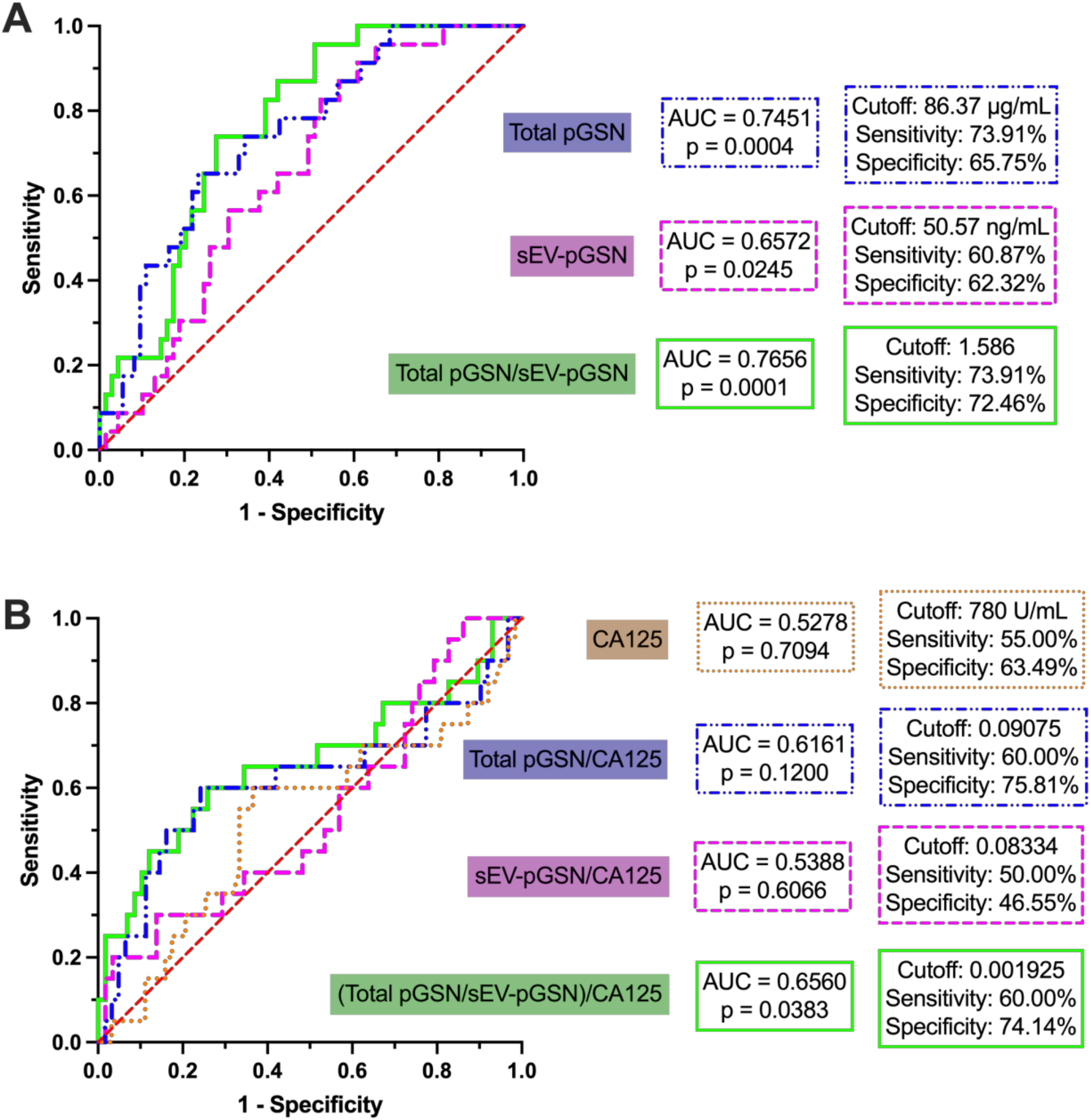
Total pGSN/sEV-pGSN outperforms other markers in predicting chemoresistance. Receiver operating characteristic curve analysis for individual and multi-analyte biomarkers to predict PFI ≤ 12 months. **(A)** Total pGSN, sEV-pGSN, and total pGSN/sEV-pGSN. **(B)** CA125, total pGSN/CA125, sEV-pGSN/CA125, and (total pGSN/sEV-pGSN)/CA125.

As expected, the introduction of CA125 in the panel of biomarkers with total pGSN or sEV-pGSN alone did not improve the ability to predict chemoresistance (PFI 6 months: Supplementary Figure 3B, PFI 12 months: Figure 3B). While the ROC analysis with the combination of all three markers did provide statistically significant results (AUC = 0.6560, p = 0.0383, Figure 3B), the test performance was not as robust as when C125 was not included, strengthening the observation that CA125 has no clinical utility in predicting EOC chemoresistance prior to the start of treatment.

## Discussion

Two of the biggest challenges in managing metastatic EOC are late-stage presentation and frequent development of resistance to chemotherapy. Being able to predict who will be at risk to develop resistant disease to common first line treatments, such as the current standard of care of platinum and taxane combination, would assist physicians in determining which course of treatment to use. Ultimately, knowing if a patient won’t respond to a cytotoxic treatment ahead of time will prevent unnecessary harm to the individual undergoing the therapy. In this study, we compared CA125, the commonly used biomarker in EOC, to total pGSN and sEV-pGSN in their ability to predict subsequent chemoresponsiveness. We found that CA125 is unable to predict chemoresponsiveness prior to treatment, while total pGSN/sEV-pGSN performed the best in this prediction. The addition of sEV-pGSN increased the test specificity that total pGSN alone lacked. A prognostic predictive biomarker of chemoresponsiveness is particularly important given current biomarkers, such as CA125, are only used in patient screening and treatment monitoring (5,6).

In addition to its use as a diagnostic biomarker for EOC, CA125 is also used to monitor the response of patients to treatment. While this has proven to be effective, serum CA125 measured prior to chemotherapy is ineffective in predicting survival (27,28). Our results align well with these previous studies, where CA125 could not predict the time between completion of first line treatment and recurrence, the metric which determines a patient’s chemoresponsiveness and eventual prognosis. It is of special importance to find non-invasive biomarkers that allow for accurate prediction of chemoresponsiveness and prognosis.

sEVs and their cargo are an emerging source of biomarkers that can be obtained from liquid biopsies, such as plasma and urine (29–32). In the context of cancer, plasma-derived sEVs that originate from the tumour cells carry the molecular signature of these malignant cells (10), thus offering an opportunity to detect markers of malignancy or cancer progression. While some research has evaluated biomarkers for monitoring treatment response, there still exists a gap in knowledge of sEV-markers that could predict chemoresistance prior to the start of treatment which will significantly impact treatment planning and patient risk stratifications.

Most groups that have investigated mechanisms by which sEV cargo promote chemoresistance within the EOC tumour microenvironment *in vitro* (11–13,33,34) have not translated these findings to a clinical context. Previous work in our laboratory highlighted the relationship of pGSN with chemoresistance in EOC cell lines and demonstrated an increase in sEV-pGSN in chemoresistant compared to chemosensitive cells (14). To address the above-mentioned gap in knowledge, we evaluated whether sEV-pGSN in plasma samples collected before primary treatment predicts the subsequent response to treatment. Excitingly, our results appear to support this finding and suggests that elevated sEV-pGSN secretion from chemoresistant EOC tumour cells is reflected in chemoresistant patients. This offers an interesting opportunity to use sEV-pGSN as a biomarker in a clinical context.

Compared to the current standard biomarker of EOC that cannot be used to predict chemoresponsiveness (8), the ratio of total pGSN/sEV-pGSN could predict chemoresistance prior to treatment with a sensitivity and specificity of 73.9% and 72.5%, respectively. The combination of sEV-pGSN with total pGSN as a multi-analytic biomarker outperformed total pGSN alone by increasing its test specificity. This highlights the clinical utility of sEV biomarkers and multi-analyte biomarkers. Validation of these findings in a larger cohort of EOC patients will be necessary to prove clinical feasibility of this biomarker. If physicians can predict a patient’s response to chemotherapy, this could help to inform their treatment strategy. Improving the likelihood that a patient will respond to the chosen therapy will undoubtably improve survival outcomes of EOC patients.

While this work provides an interesting proof of concept for evaluating pGSN as a biomarker for predicting EOC patients’ response to chemotherapy, there is an important limitation that will need to be addressed in future work. The cohort of patients included in this study are primarily of the high grade serous histologic subtype and in FIGO stage III. We cannot conclude whether pGSN is a clinically relevant biomarker of chemoresistance in other EOC subtypes or in earlier FIGO stages. Future work in which there is an adequate representation of histopathological subtypes and FIGO stages is necessary. Given the biological difference between serous and non-serous EOC, a larger cohort with better representation of histologic subtypes will allow us to determine the relevance of pGSN and sEV-pGSN are applicable as biomarkers of chemoresistance across histologic subtypes of EOC.

With much research emerging to identify biomarkers of ovarian cancer, it is important to take this work past the bench and onto the bedside. Screening for identifiers of poor response to chemotherapy at the time of diagnosis would save valuable time in aggressive treatment in EOC patients. Once these findings are validated in the larger context of histopathological subtypes and FIGO stages, pursuit of clinical trials to interrogate pGSN as a predictor of chemoresistance is necessary. The finding that multi-analytic marker total pGSN/sEV-pGSN at the time of diagnosis predicts chemoresistance highlights the importance of using of multi-analytic biomarkers to maximize test performance, to inform clinicians’ therapeutic approach to EOC, and to improve patient outcomes.

## Materials and Methods

### Plasma samples

The 96 plasma samples used in this study were obtained from the Banque cancer de l’ovaire, Centre de recherche du CHUM (CRCHUM), in Montreal, Quebec, Canada. These samples were collected between the years of 1992-2012 from individuals diagnosed with EOC and *before* any treatment (chemotherapy or radiotherapy). Gynecologic-oncology pathologists reviewed patient tumour samples to determine histopathological subtype and stage, as per the FIGO criteria. CA125 was measured in the clinic at the time of sample collection. Patient demographics, including age, histopathological subtype, FIGO stage, and PFI are described in Table 1. All patients underwent an initial surgical debulking followed by chemotherapy. Progression free interval (PFI) is defined as the time between diagnosis and recurrence.

### sEV isolation from plasma samples

To isolate sEVs from patient plasma samples, ExoQuick ULTRA EV Isolation System (System Biosciences, cat # EQULTRA-20A-1) was used and performed as per the manufacturer’s protocol (35). 40uL of plasma was mixed with 500uL of PBS. The sEVs were isolated using 100µL of ExoQuick reagent. The sEV depleted plasma was saved for western blot analysis. Isolated sEVs were resuspended in 500µL of 0.1µm-filtered PBS. They were subsequently divided for downstream analysis (western blot, nanoparticle tracking analysis, ELISA) and stored at -80°C (Supplementary Figure 4A - C).

### Nanoparticle tracking analysis

The concentration and size distribution of isolated sEVs were measured by nanoparticle tracking analysis (NTA). The ZetaView PMX110 Multiple Parameter Particle Tracking Analyzer (Particle Metrix, Meerbusch, Germany) was used in *Size Mode*, as previously described (36). See Supplementary Figure 4C.

### Protein extraction, quantification, and western blot

Membranes of resuspended sEVs were disrupted by sonication. Protein content from each sample was quantified using the *DC* Protein Assay (BioRad, cat #5000116). Equivalent amounts of protein (10µg) from sEVs and a positive control (endometrioid EOC cell line: A2780cp) were prepared by adding lysis buffer (Roche, cat # 04719956001) and Laemmli sample buffer (BioRad, cat # 1610737) and then boiled for 4 minutes. Samples were loaded into 12% acrylamide gels. Proteins were separated using electrophoresis (100V for 30 minutes, 120V for 90 minutes) and then transferred onto nitrocellulose membranes (110V for 90 minutes). Protein migration was assessed using Ponceau-S staining. Membranes were blocked using 5% skim milk prepared in Tris-buffered saline-Tween (TBS-T) for 1 hour. Membranes were incubated in primary antibody solutions (Rabbit polyclonal CD9; System Biosciences, cat # EXOAB-CD9A-

1. Mouse monoclonal CD63; Abcam, cat # ab193349. Rabbit polyclonal CD81; System Biosciences, cat # ECOAB-CD81A-1. Rabbit monoclonal GAPDH; Abcam, cat # ab181602. Rabbit monoclonal calnexin; Abcam, cat # ab133615) for approximately 18 hours, washed twice in TBS-T for 5 minutes, followed by incubations with the appropriate secondary antibody (Goat Anti-Mouse IgG (H + L)-HRP Conjugate; BioRad, cat # 1706516. Goat Anti-Rabbit IgG (H + L)-HRP Conjugate; BioRad, cat # 1706515) for 1 hour and final membrane washings in TBS-T 3 times for 15 minutes each. ECL Prime Western Blotting Detection Reagent (Amersham, cat #RPN2124) was used to visualize protein bands with the BioRad ChemiDoc MP. See Supplementary Figure 4A.

### Immunoelectron microscopy (iEM)

Isolated sEVs were pelleted by ultracentrifugation (100,000g for 90 minutes) and fixed as previously described (37). The iEM protocol is previously described by Asare-Werehene *et al*.

(14) using a monoclonal anti-pGSN antibody (ABGENT, cat # AM1936a). See Figure 4B.

### Enzyme-linked immunosorbent assay

To measure both total circulating and sEV-specific pGSN, enzyme-linked immunosorbent assay (ELISA) was performed. The human soluble plasma gelsolin sandwich ELISA kit from Aviscera Bioscience Inc. (SK00384-01) was used as per the manufacturer’s instructions. Plasma samples were prepared using a 1/15000 dilution, while resuspended sEVs were prepared with a 1/5 dilution. Concentrations were measured in singlet, with the blank OD being subtracted from each sample reading. Total pGSN concentrations are reported in µg/mL while sEV-pGSN concentrations are reported in ng/mL.

### Statistical analysis

All statistical analyses were performed using GraphPad Prism version 9.4.1. To compare biomarker means between chemosensitive and chemoresistant groups, Student t-test or Mann-Whitney U-test were used as appropriate. ROC analysis was used to compare clinical test performances of biomarkers.

## Data Availability

All data produced in the present study are available upon reasonable request to the authors.

## Declarations

### Ethics approval and consent to participate

This study was approved by both the Centre hospitalier de l’Uniersité de Montréal (CHUM) ethics committee (IRB approval number: BD 04-002) and the Ottawa Health Science Network Research Ethics Board (IRB approval number: OHSN-REB 1999540-01H). All work followed appropriate guidelines and all patients provided written informed consent.

### Consent for publication

Not applicable.

### Availability of data and materials

The datasets used and/or analysed during the current study are available from the corresponding author on reasonable request.

### Authors’ contributions

MAW, EG and BKT conceived and designed the study. EG performed sEV isolation and ELISA. sEV characterization was performed and analyzed by EG and DB. sEV fixation, immunoelectron microscopy and analysis were performed by EG, MAW and AR. AMMM and EC provided plasma samples. TL informed analysis and interpretation of clinical data. Statistical analyses were done by EG and MAW. EG wrote the paper with scientific feedback from all authors.

## Acknowledgements

Banking of plasma samples was supported by the Banque de tissus et de données of the Réseau de recherche sur le cancer (RRCancer) du Fonds de recherche du Québec – Santé (FRQS), associated with the Canadian Tissue Repository Network (CTRNet). AMMM is a researcher of the CRHUM, which receives support from the FRQS.

