## Supplementary material for "Predicting chemoresponsiveness in epithelial ovarian cancer patients using circulating small extracellular vesicle-derived plasma gelsolin": Table 1.

**Table 1.** Patient characteristics.

| Characteristic | Number (n=96) | % |
| --- | --- | --- |
| Age (range: 36-82, average 61) < 61  ≥ 61 | 47  49 | 49  51 |
| Histopathologic Subtype  High-grade serous  Low-grade serous  Undefined | 69  1  26 | 72  1  27 |
| FIGO Stage  1  2  3  4 | 8  10  67  11 | 8.3  10.4  69.8  11.5 |
| Progression Free Interval  ≤ 6 months  > 6 months  ≤ 12 months  > 12 months | 9  87  23  73 | 9  91  24  76 |
