## Supplementary Figures. for "Predicting chemoresponsiveness in epithelial ovarian cancer patients using circulating small extracellular vesicle-derived plasma gelsolin"

### Slide 1
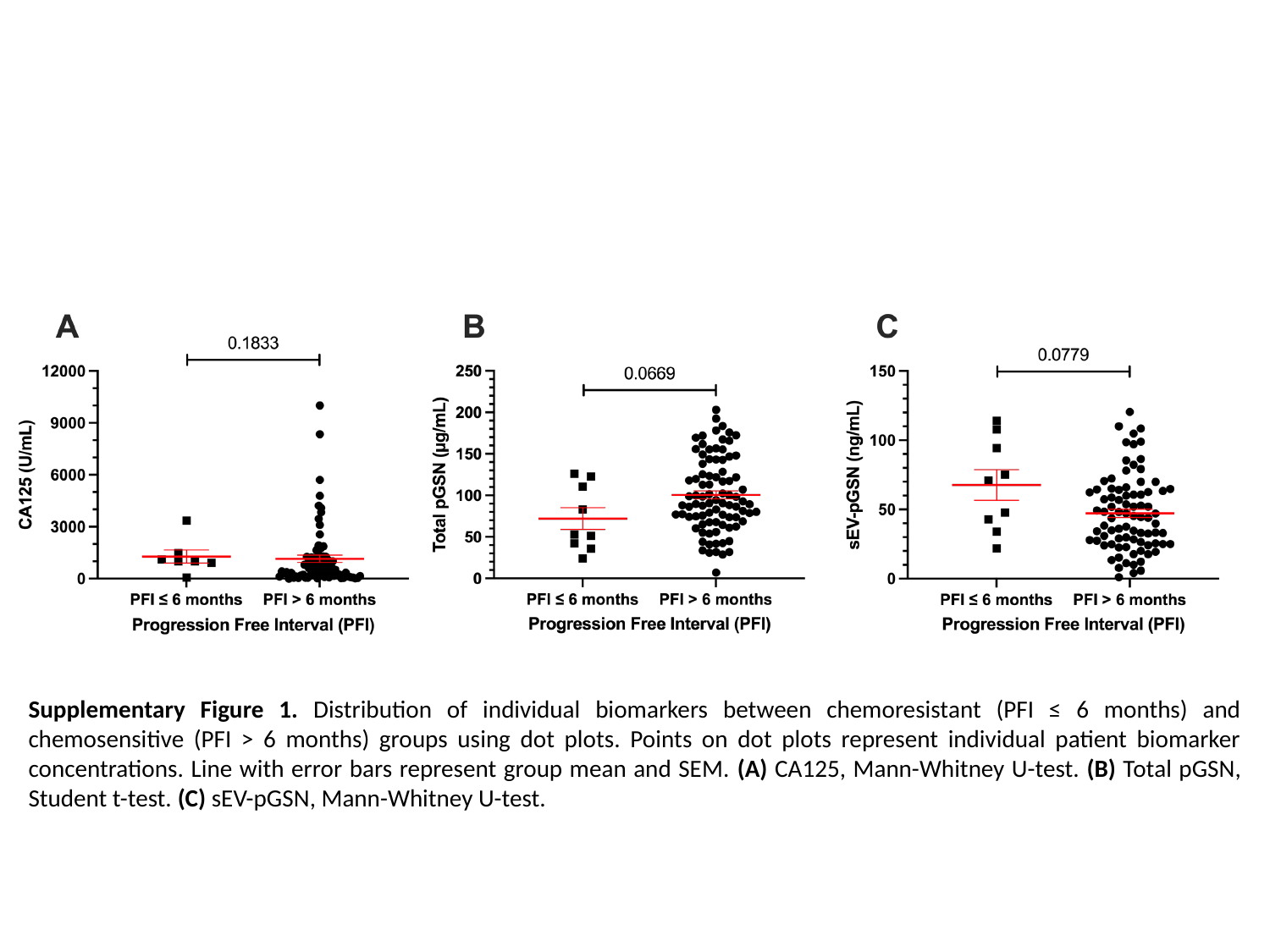

Supplementary Figure 1. Distribution of individual biomarkers between chemoresistant (PFI ≤ 6 months) and chemosensitive (PFI > 6 months) groups using dot plots. Points on dot plots represent individual patient biomarker concentrations. Line with error bars represent group mean and SEM. (A) CA125, Mann-Whitney U-test. (B) Total pGSN, Student t-test. (C) sEV-pGSN, Mann-Whitney U-test.

### Slide 2
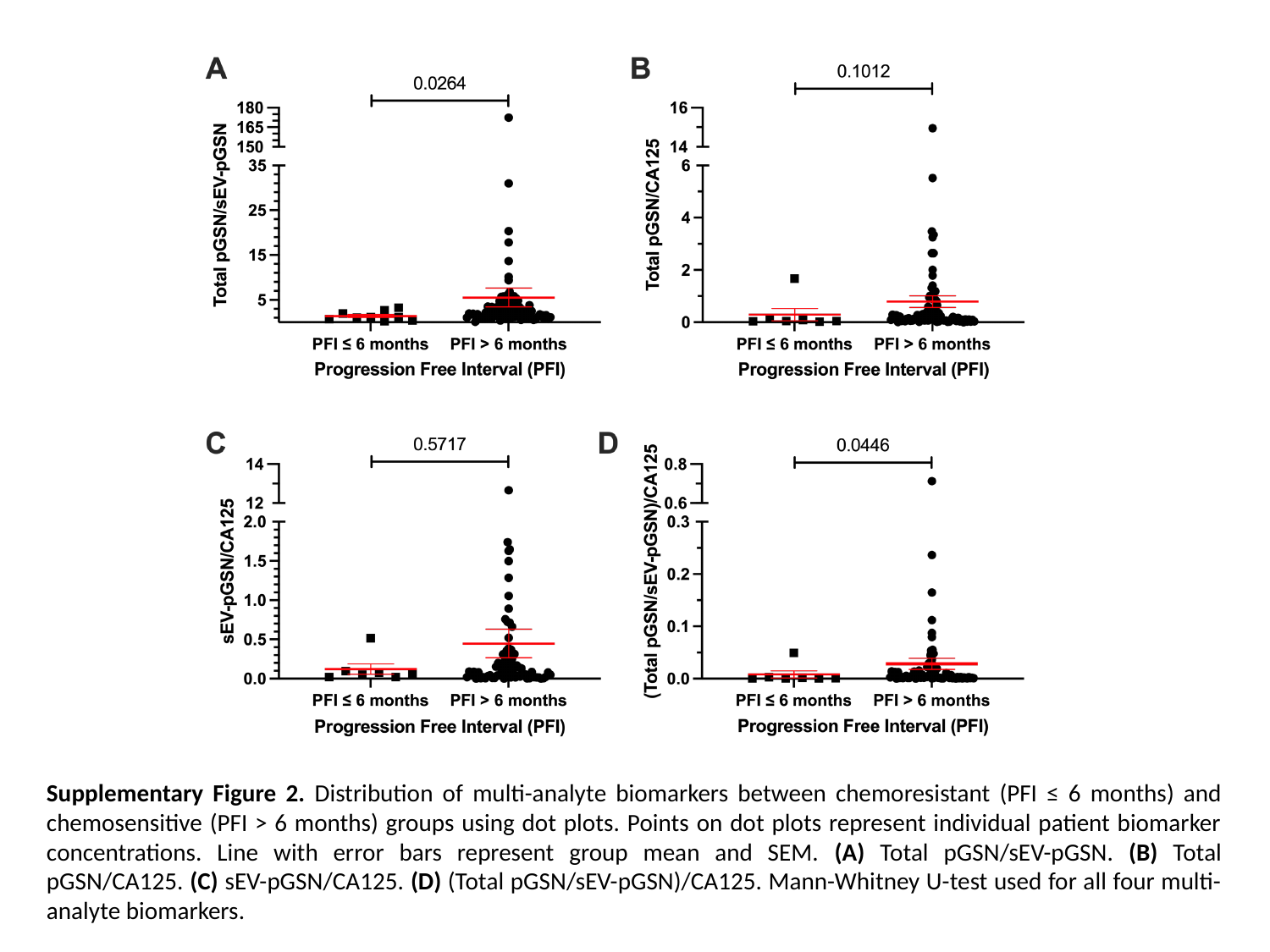

Supplementary Figure 2. Distribution of multi-analyte biomarkers between chemoresistant (PFI ≤ 6 months) and chemosensitive (PFI > 6 months) groups using dot plots. Points on dot plots represent individual patient biomarker concentrations. Line with error bars represent group mean and SEM. (A) Total pGSN/sEV-pGSN. (B) Total pGSN/CA125. (C) sEV-pGSN/CA125. (D) (Total pGSN/sEV-pGSN)/CA125. Mann-Whitney U-test used for all four multi-analyte biomarkers.

### Slide 3
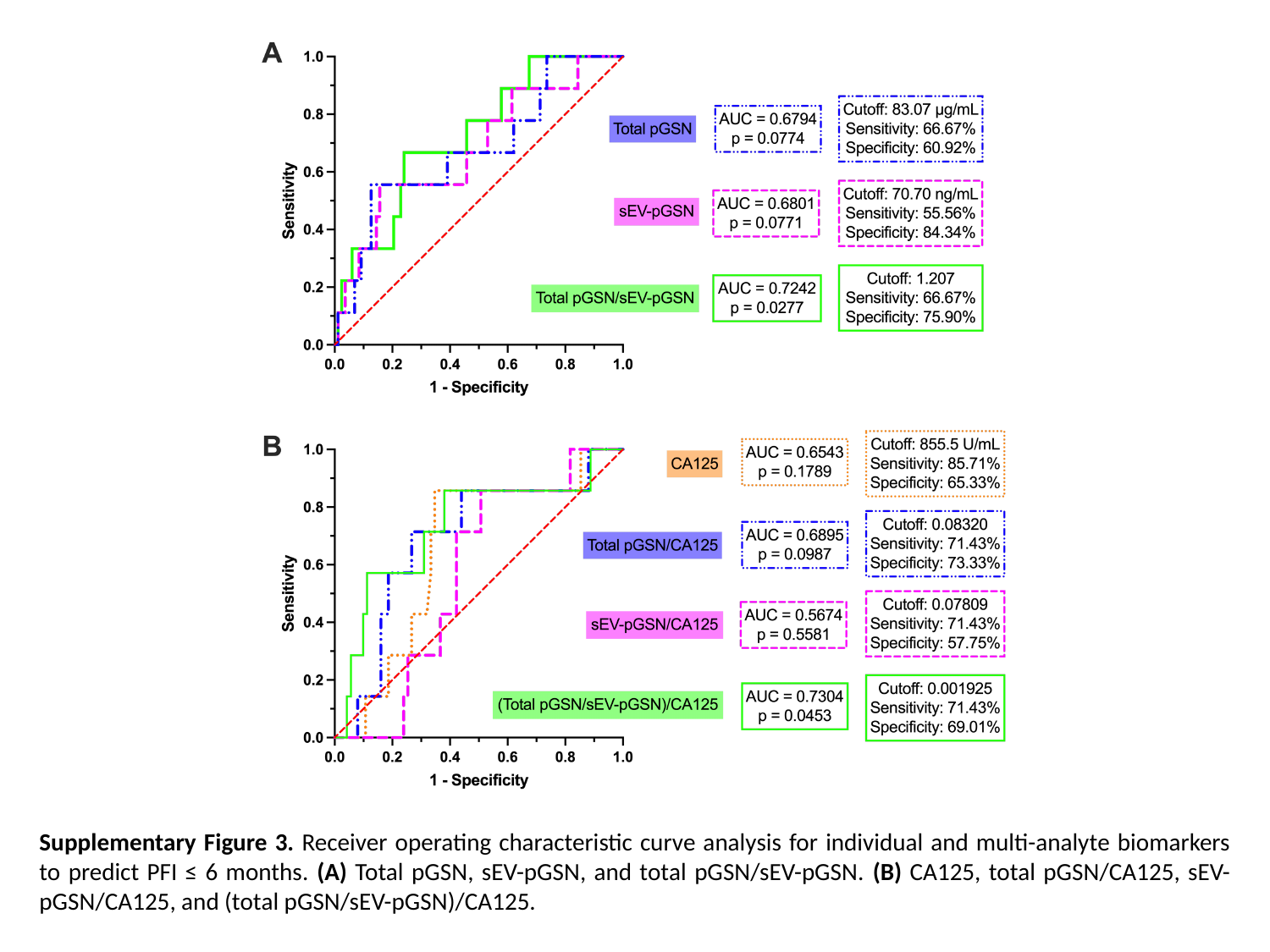

Supplementary Figure 3. Receiver operating characteristic curve analysis for individual and multi-analyte biomarkers to predict PFI ≤ 6 months. (A) Total pGSN, sEV-pGSN, and total pGSN/sEV-pGSN. (B) CA125, total pGSN/CA125, sEV-pGSN/CA125, and (total pGSN/sEV-pGSN)/CA125.

### Slide 4
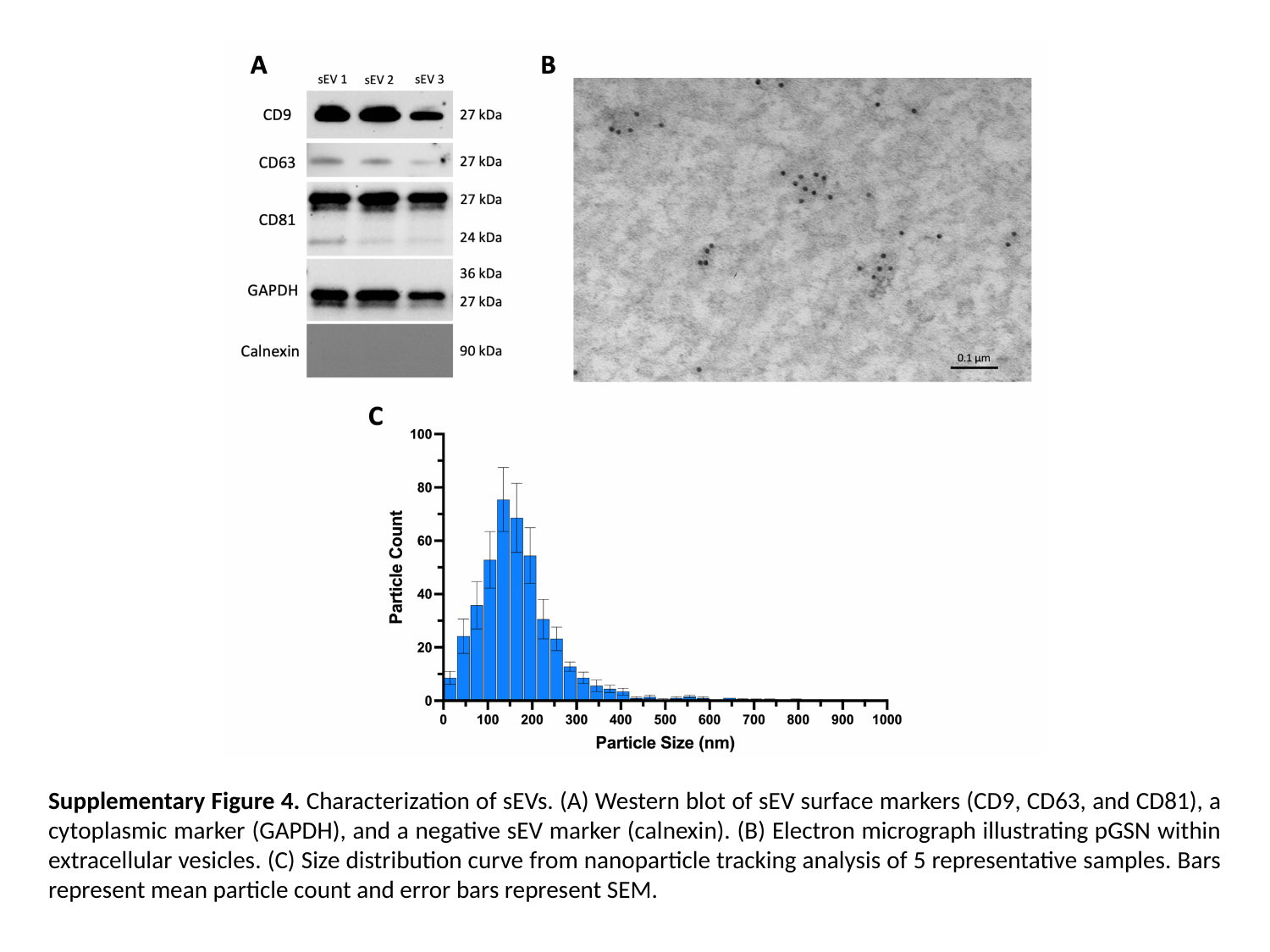

Supplementary Figure 4. Characterization of sEVs. (A) Western blot of sEV surface markers (CD9, CD63, and CD81), a cytoplasmic marker (GAPDH), and a negative sEV marker (calnexin). (B) Electron micrograph illustrating pGSN within extracellular vesicles. (C) Size distribution curve from nanoparticle tracking analysis of 5 representative samples. Bars represent mean particle count and error bars represent SEM.
